# Statistical Relationship Between Wastewater Data and Case Notifications for COVID-19 Surveillance in the United States, 2020-2023: A Bayesian Hierarchical Model

**DOI:** 10.1101/2024.10.20.24315840

**Authors:** Masahiko Haraguchi, Fayette Klaassen, Ted Cohen, Joshua A. Salomon, Nicolas A. Menzies

**Affiliations:** The Department of Global Health and Population, Harvard T.H. Chan School of Public Health, 677 Huntington Avenue, Boston MA, USA; Department of Epidemiology of Microbial Diseases and Public Health Modeling Unit, Yale School of Public Health, 60 College St, New Haven CT, USA; Department of Health Policy, Stanford University School of Medicine, 1265 Welch Road, Stanford CA, USA

**Keywords:** wastewater epidemiology, COVID-19 surveillance, environmental surveillance, SARS-CoV-2, Bayesian modeling, the pandemic

## Abstract

During the COVID-19 pandemic a number of jurisdictions in the United States began to regularly report levels of SARS-CoV-2 in wastewater for use as a proxy for SARS-CoV-2 incidence. Despite the promise of this approach for improving situational awareness, the degree to which viral levels in wastewater track with other outcome data has varied, and better evidence is needed to understand the situations in which wastewater surveillance tracks closely with traditional surveillance data. In this study, we quantified the relationship between wastewater data and traditional case-based surveillance data for multiple jurisdictions. To do so, we collated data on wastewater SARS-CoV-2 RNA levels and COVID-19 case reports from July 2020 to March 2023, and employed Bayesian hierarchical regression modeling to estimate the statistical relationship between wastewater data and reported cases, allowing for variation in this relationship across counties. We compared different model structural approaches and assessed how the strength of the estimated relationships varied between settings and over time. These analyses revealed a strong positive relationship between wastewater data and COVID-19 cases for the majority of locations, with a median correlation coefficient between observed and predicted cases of 0.904 (interquartile range 0.823 – 0.943). Across locations, the COVID-19 case rate associated with a given level of wastewater SARS-CoV-2 RNA concentration declined over the study period. Counties with higher population size and of higher levels of urbanicity had stronger concordance between wastewater data and COVID-19 cases. Ideally, use of wastewater data for decision-making should be based on an understanding of their local historical performance.

## Introduction

The practice of employing wastewater data to track pathogens has gained significant interest as an innovative method of infectious disease surveillance, with nearly 80% of the United States population connected to public wastewater systems (U.S. Government Accountability Office, 2022; Yu et al., 2023). During the COVID-19 crisis, local health agencies started to gather and report data on COVID-19 concentrations in wastewater, using the trends in these data as an indirect indicator of SARS-CoV-2 transmission patterns.

Tracking the presence and concentration of pathogens in wastewater, a passive method of environmental surveillance, has been used for several decades to track infectious diseases, such as polio (Matrajt et al., 2018), gastroenteritis (Kazama et al., 2017), hepatitis E (Alfonsi et al., 2018), and acute diarrhea (Prevost et al., 2015), among others. However, the COVID-19 pandemic accelerated interest in this approach worldwide (Naughton et al., 2023), as it offers multiple benefits as a complement to traditional surveillance systems. First, it can serve as an early warning system for SARS-CoV-2 transmission, sensitive to asymptomatic and presymptomatic cases four to ten days prior to clinical testing signals (Wu et al., 2022). Second, it can monitor community-level transmission when implemented at downstream locations such as wastewater treatment plants (McClary-Gutierrez et al., 2021). The fact that it does not require individual testing circumvents the challenges created by variable supply of and demand for COVID-19 diagnostic testing (as has been observed over the pandemic), and the decline in reporting of test results (National Academies of Sciences & Medicine, 2023). Third, wastewater surveillance programs with specific methods can track virus variants in the early stage of their evolution, allowing for early identification of emerging variants (Jahn et al., 2022; Karthikeyan et al., 2022; McClary-Gutierrez et al., 2021). Lastly, it provides an early indicator of epidemiological changes (as compared to hospitalization and death data), so that mitigation and other response measures can be deployed more rapidly. With these advantages, tracking wastewater COVID-19 data is a potentially powerful tool for COVID-19 surveillance.

Despite the potential advantages of SARS-CoV-2 wastewater surveillance, significant challenges remain. Existing studies have generally considered the relationship between reported cases and wastewater metrics at a limited number of locations, such as university dorms (Kotay et al., 2022), nursing homes (Davó et al., 2021), university campuses (Karthikeyan et al., 2021), and municipalities (e.g., Oklahoma City, Oklahoma (Kuhn et al., 2022) and Louisville, Kentucky (Klaassen et al., 2024)). While these studies provide valuable insights, they often do not fully account for heterogeneities across different geographical locations. Furthermore, the time periods covered by existing studies are limited. For example, Xiao et al. (2022) associated clinical case data with wastewater data within three Massachusetts counties from March 2020 through May 2021. Similarly, Weidhaas et al. (2021) analyzed nine weeks of wastewater and COVID-19 case data related to ten wastewater treatment facilities in Utah. Notably, reported cases themselves are not a perfect indicator for true infection situations, given that they depend on various factors such as testing availability and access as well as the number of asymptomatic infections. As such, while quantifying the relationship between reported cases and wastewater metrics provides valuable insights, it should be noted as the next-best alternative. Studies that compare the performance of COVID-19 wastewater surveillance data across sewersheds over extended time frames remain scarce (Dai et al., 2024). Such research is valuable for establishing the statistical basis for real-time trend analysis, and describing the conditions under which wastewater surveillance performs well.

This study explored the quantitative relationship between SARS-CoV-2 wastewater surveillance data and reported COVID-19 diagnoses across multiple sewer-sheds over the initial years of the COVID-19 pandemic. Using weekly-aggregated wastewater and case report data for 107 U.S. counties over 2020-2023, we employed Bayesian hierarchical modeling to establish the statistical relationship between these two data sources, describe changes in these relationships over time and across locations, and identify how the strength of these relationships varied systematically by county characteristics.

## Data and Methods

### Data sources

For the study period July 1, 2020 to March 1, 2023, we collated county-level SARS-CoV-2 wastewater surveillance data reported by Biobot Analytics, including 254 counties covering approximately 30% of the U.S. population. These data represent RNA copies per milliliter, normalized by the concentration of pepper mild mottle virus (PMMoV) to correct for variability in fecal content, which is influenced by environmental factors such as stormwater (Duvallet et al., 2022). Surveillance data on county-level weekly COVID-19 reported case totals were extracted from the COVID-19 data repository in the Center for Systems Science and Engineering at Johns Hopkins University.

Due to varied implementation of wastewater surveillance operations, wastewater data were not available for all county-weeks. We restricted the analysis to counties with a minimum of 50 weeks of available wastewater data during the study period, which resulted in 107 counties being included, covering a range of geographic areas within the United States, and with periods of data incompleteness for the majority of counties. We grouped counties into six ordinal urbanicity categories as defined by the National Center for Health Statistics (NCHS). Categories ranged from NCHS category 1 (large central metro) as the most urban to NCHS category 6 (non-core) as the most rural. Fig1 shows the geographic distribution of the 107 counties included in the analysis, coded by NCHS urbanicity category.

**Figure 1:**
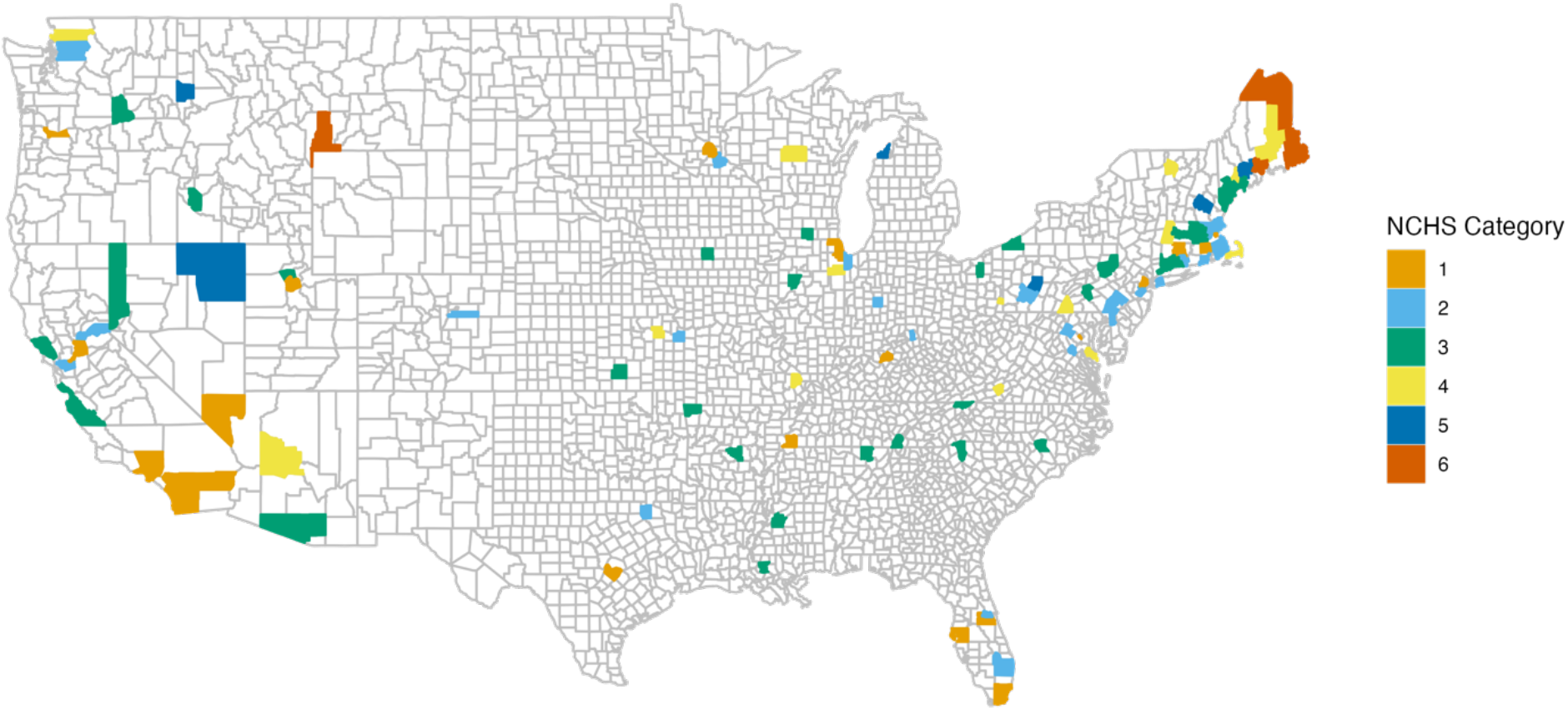
This shows geographic location and NCHS urbanicity category for the 107 counties included in the analysis.

### Hierarchical regression models

Using the wastewater and COVID-19 case data, we constructed hierarchical Bayesian regression models to capture key features of each data source, allowing for differences in the estimated relationship between these data over time and between modeled counties. Accounting for temporal variations is vital as the dynamics of the pandemic changed over time due to factors such as the emergence of new variants, public health interventions, vaccination rollouts, and population immunity. Also, allowing for spatial variations accounts for local differences, such as vaccination coverage, and socioeconomic and demographic characteristics. As all of them influence the relationship between wastewater metrics and reported cases, a model that can account for the complex and evolving nature of the pandemic is critical. Namely, we modelled the weekly COVID-19 case reports for each county using a negative binomial likelihood, allowing for over-dispersion in these data:

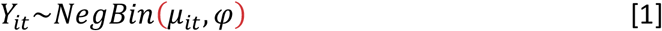

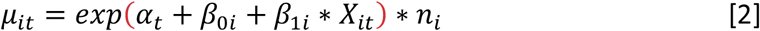

In Equation 1, *Y_it_* represents reported COVID-19 cases for county *i* and week *t.* We used an alternative parameterization of the negative binomial in which *μ_it_* parameterizes the mean of the likelihood and *φ* parameterizes the extra-Poisson variation. The mean was specified as a function of *α_t_*, a time-varying coefficient given a random-walk prior; *β*_0*i*_, a county-specific intercept; and *β*_1*i*_, a county-specific slope term applied to *X_it_*, the demeaned wastewater COVID-19 concentration value for each county and week (Equation 2). Both *β*_0*i*_ and *β*_1*i*_ were specified as random effects to pool information across counties. *n_i_* represented the population of each country, as reported by the US Census Bureau. We fit this model to the COVID-19 case and wastewater data using the RStan package in R (Stan Development Team, 2022). Additional details on the specification of this regression model, including prior distributions, are provided in the Appendix.

### Analysis of fitted models

We used several approaches to assess model fit. First, we visually compared the COVID-19 case time series for each county to the fitted values from the regression model. Second, we calculated a quantitative measure of model fit: the Median Absolute Deviation (MAD). We estimated this value for each county and used them to describe the overall level of model fit and how this varied across counties. We used the MAD to investigate whether the strength of estimated relationships differed systematically as a function of county characteristics (urbanicity, population size), examining these relationships visually and via univariable and multivariable regression models. Finally, we calculated the correlation coefficient between modelled values and raw case totals as a simple summary measure of model fit.

### Coefficient estimates

We used the fitted values of *α_t_* (the temporal trend in the regression model) to understand how the relationship between wastewater and COVID-19 case totals varied over the study period (July 2020 to March 2023). We used the fitted values of *β*_0*i*_ and *β*_1*i*_ to understand how the relationship between wastewater concentration and COVID-19 case totals varied within each county.

## Results

### Fitted relationship between COVID-19 cases and wastewater concentration

For the majority of modelled locations, we estimated a relationship between wastewater concentration and weekly COVID-19 cases, indicating that wastewater concentration serve as a useful predictor of case trends. Figure 2 shows the temporal tend in reported COVID-19 cases and fitted model estimates for each of six example counties, representing a range in terms of urbanicity and population size, which are often correlated but not identical. For each of these example countries the fitted model values (blue symbols) follow the empirical case data (black symbols) closely, with occasional deviations (e.g., 2021 estimates for Monterey County, CA). Figures for other counties are available in the Appendix. We also calculated correlation coefficients comparing observed and predicted values COVID-19 case counts. Across counties the median of these correlation coefficients was 0.904, with an inter-quartile range of 0.823 to 0.943. Twenty-three counties (21.5% of the sample) had correlation coefficients below 0.8, and three had values below 0.6.

**Figure 2:**
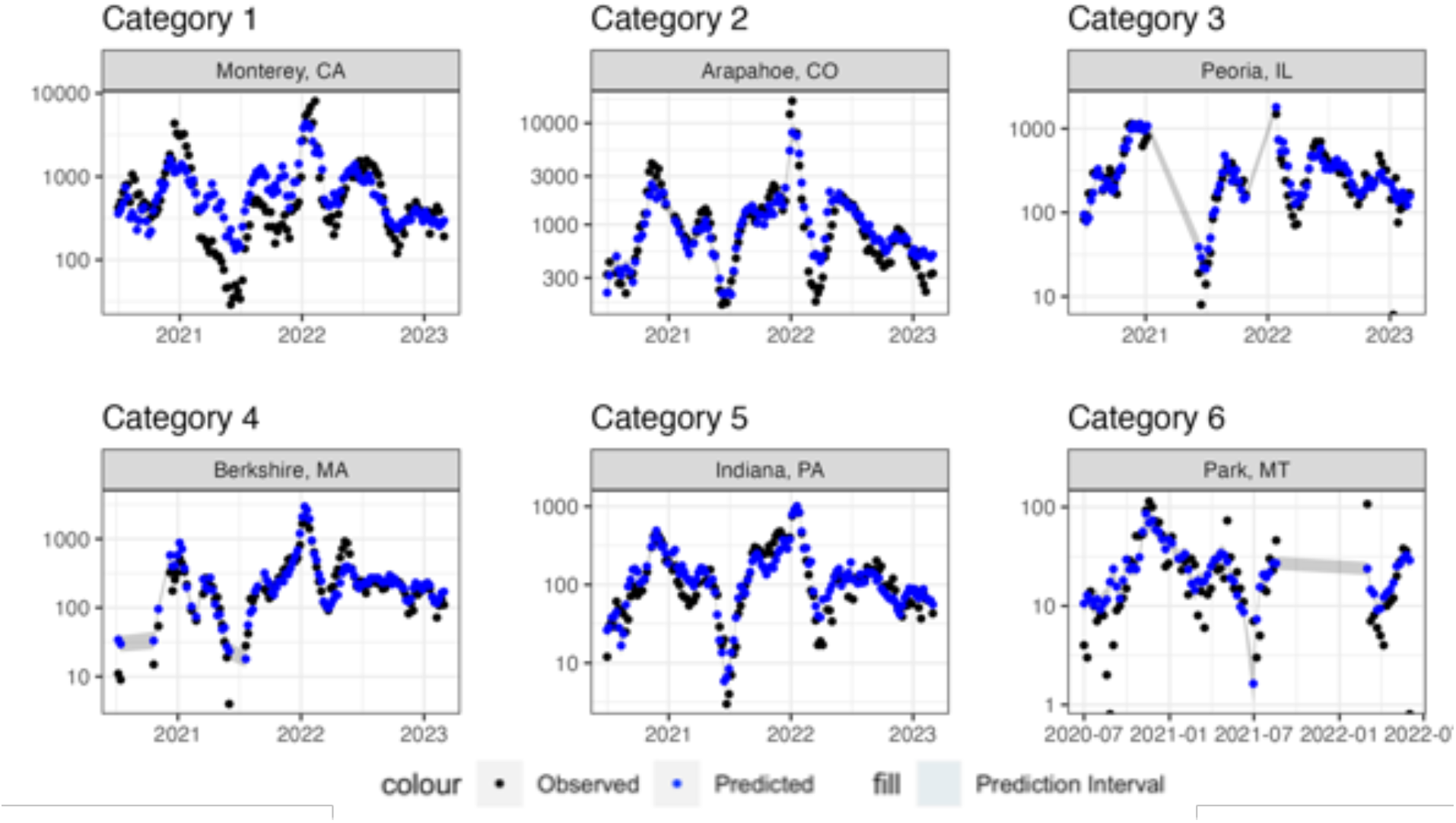
Comparison of observed and predicted COVD-19 case counts for a select group of counties within each urban-rural category as defined by the National Center for Health Statistics (NCHS).

### Systematic differences in model fit across counties

To further evaluate how the model performed in each county we calculated the MAD, for which smaller values indicate better model fit. For MAD, the median value was 0.259, with an interquartile range of 0.201 to 0.301. When we compared MAD to country population size(Figure 3-a), we found that model fit was better for counties with higher population numbers (p=<0.001).

**Figure 3:**
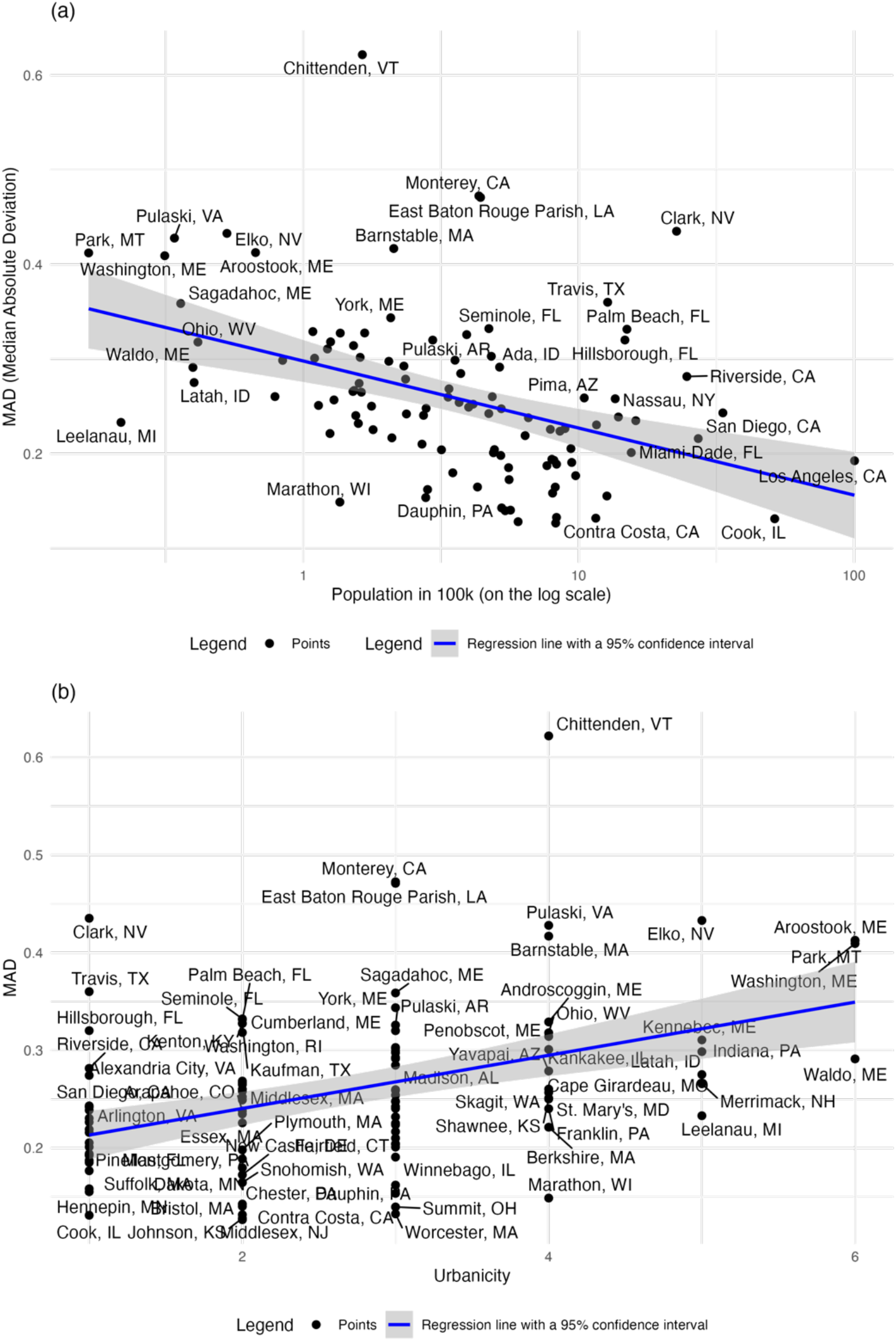
Panel A shows how the quality of model fit (MAD) varies with county population size. Panel B shows how the quality of model fit (MAD) varies with country urbanicity category.

Furthermore, we found that MAD was associated with urbanicity (Figure 3-b), with more urban counties (Categories 1, 2, and 3) having lower MAD (p=<0.001) and therefore better model fit. More rural counties had poorer model fits (higher values of MAD), with the exception of Chittenden VT.

When we fit a multivariable regression model including both logged population and urbanicity category as predictors we found both coefficients to have the same sign as in the univariate analyses but were no longer significant (population size: p=0.09, urbanicity: p=0.13).

### Time-trends

Figure 4 shows changes in the estimated relationship between wastewater concentration and COVID-19 case reports over the study period, quantified as the level of logged COVID-19 case totals consistent with a given wastewater concentration, shown in blue. Notably, the fluctuations in this relationship are closely associated with the significant US waves at the end of 2020 to the beginning of 2021 (Alpha wave), the summer of 2022 (Delta wave), and the beginning of 2022 (Omicron wave). During these periods, the ratio of COVID-19 cases to wastewater concentration is relatively high, as compared to the months before and afterwards. After early 2022 the estimated trend shows a progressive decline.

**Figure 4:**
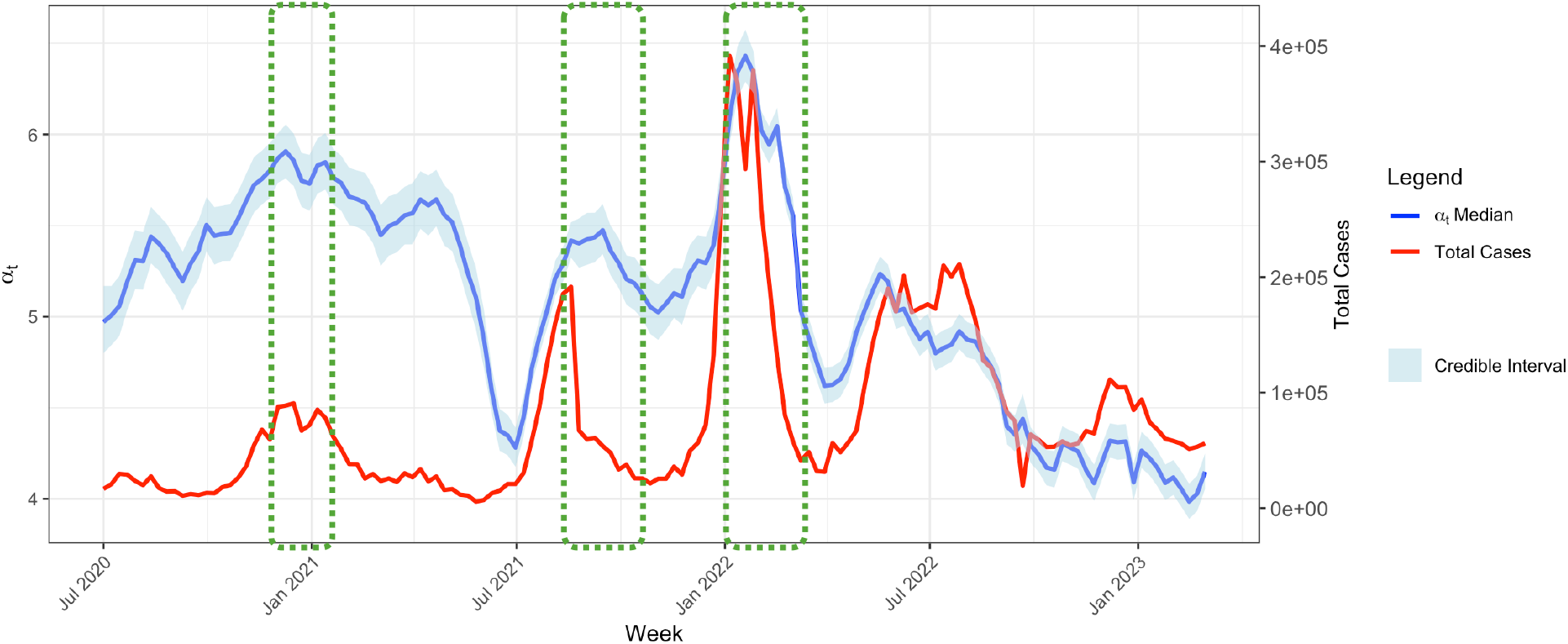
Time series of the time-varying coefficient (*α_t_*) alongside the aggregated case counts across counties from July 2020 to February 2023.

### Differences across counties

Figure 5 shows fitted estimates for how changes in wastewater concentration values are associated with changes in COVID-19 cases for each county. As expected, the slopes shown in Figure 5 are positive (indicating that an increase in wastewater concentration was associated with an increase in reported cases), and relatively consistent across counties. In all cases the slope of these lines was estimated to be less than 1.0 (median: 0.551, IQR: 0.447 to 0.632), indicating that that fitted relationship between wastewater concentration and COVID-19 case totals is less than proportional (e.g., a 50% increase in wastewater concentration is associated with a <50% increase in case totals).

**Figure 5:**
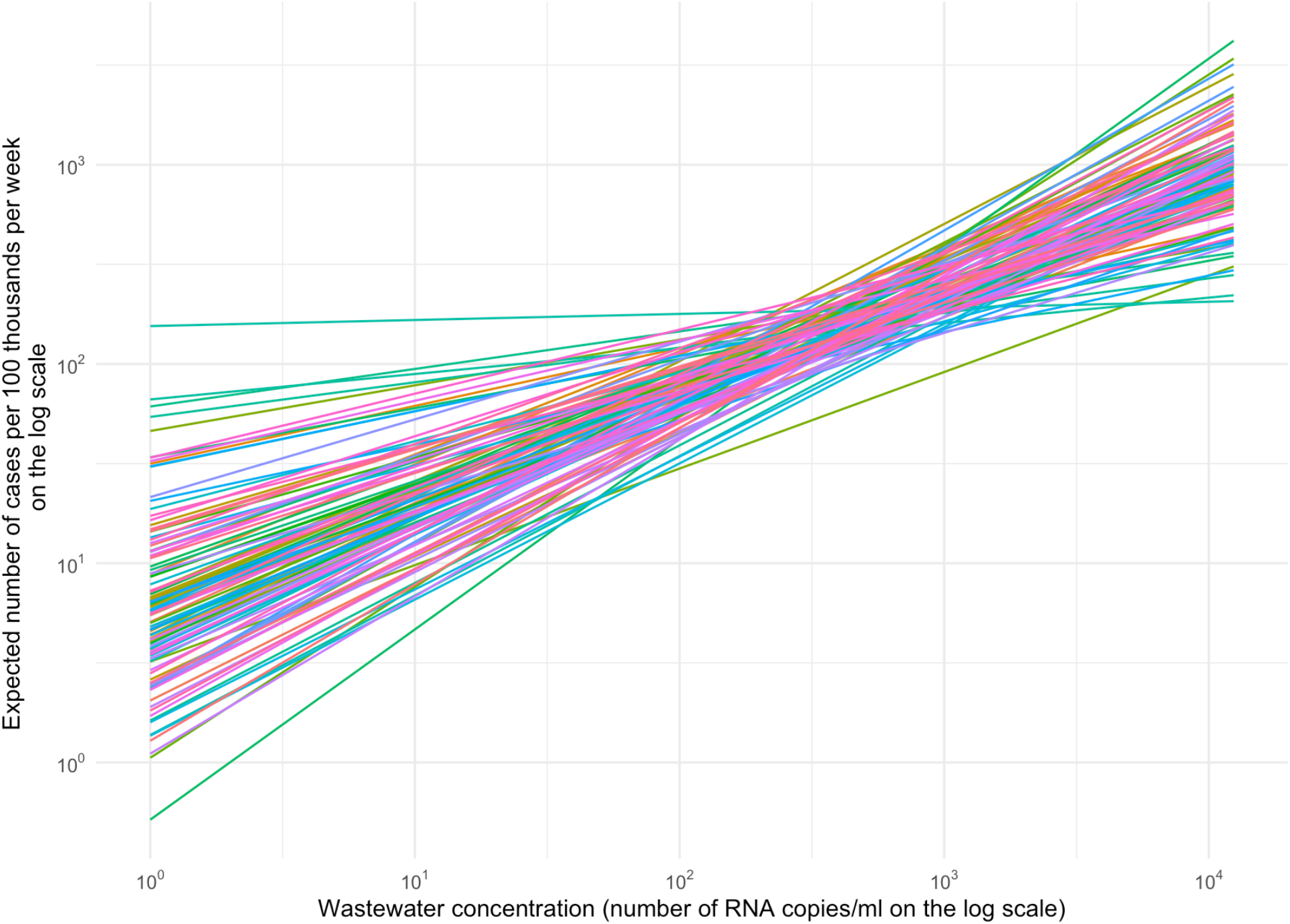
Estimated relationship between wastewater concentration and mean weekly cases per 100 thousand people for each county.

## Discussion

Wastewater data have been used extensively during the SARS-CoV-2 pandemic to monitor disease trends and provide early evidence of rising community transmission. However, there is limited information on the statistical relationship between wastewater metrics and reported COVID-19 cases, and how this relationship varies over time and across jurisdictions (Dai et al., 2024). This knowledge is valuable for making decisions about how best to use wastewater data, and to understand the settings in which these data provide accurate information about COVID-19 case trends. In this study, we modeled the relationship between wastewater metrics and clinical cases at the county level in the United States from July 2020 to March 2023.

The results of our analysis show that models fit to wastewater data are better able to predict case counts in urban counties (based on NCHS categorization) as compared to more rural counties. This may be due to rural areas having lower levels of connection to centralized sewage systems, the source of wastewater surveillance data (National Academies of Sciences & Medicine, 2023; M. Varkila et al., 2023). We also noted a reduction in model performance among counties with smaller population sizes. This is consistent with other studies that have reported wastewater surveillance to have limited sensitivity as an early warning indicator in smaller geospatial scales (Gamage et al., 2024; Klaassen et al., 2024). We also estimated differences in the quality of model fit that were not explained by urbanicity and population size—these may relate to local differences in the coverage of wastewater surveillance, the processing of wastewater samples, or the quality of COVID-19 case reporting.

In addition to inter-county differences, the results also revealed fluctuations in the relationship between wastewater concentration and COVID-19 case totals over the course of the pandemic. Several factors could account for these temporal trends. First, viral shedding patterns among individuals developing COVID-19 in recent years likely differ from those developing disease in the early stage of the pandemic, as immunity in the population through previous infections and vaccination increased significantly throughout the pandemic (Puhach et al., 2023). Second, the transition between different dominant variants may have also influenced the dynamics of discharged RNA copies in human waste. For example, the viral and antibody dynamics are distinct between omicron and delta variants (Yang et al., 2023). Also, as mutations may affect the quantification of SARS-CoV-2 concentration in wastewater, such viral changes may need to be accounted for in estimating the relationship between wastewater levels and case notifications (Endo et al., 2023). Lastly, case reporting systems have changed over time, influencing the ratio of reported and unreported cases (Alvarez et al., 2023; Silk et al., 2023). In particular, the declining trend over the final year of the time series (i.e. a declining number of reported COVID-19 cases for a given wastewater level) likely relates to changes in COVID-19 testing and reporting practices, with a progressively smaller fraction of COVID-19 cases diagnosed and reported to public health authorities.

Designing effective wastewater surveillance systems require trade-offs among cost-effectiveness, speed, and local feasibility (Levy et al., 2023). Most current sequencing is implemented with hundreds to thousands of samples in parallel with expensive machinery and intensive investment in human resources (Levy et al., 2023). This implies that counties with fewer resources may find it difficult to finance and support the required laboratory infrastructure and human resources.

While the quality of model fit was generally good, our analyses revealed substantial variation in the utility of wastewater surveillance across counties. It is also important to note that reported case counts, which we used as a proxy for infection trends, are an imperfect measure of true incidence (M. R. Varkila et al., 2023). Ideally, a population-based survey, such as the United Kingdom’s Office for National Statistics’ COVID-19 Infection Survey, would provide a more accurate information for assessing the predictive performance of wastewater surveillance. However, without such data in the US, we rely on case reports as the best available data. Further investigation, validation, and standardized data collection frameworks are required to better understand the relationship between wastewater and epidemiological data. The incorporation of next-generation sequencing and automation of wastewater data collection processes could enhance the effectiveness of wastewater surveillance (Iwamoto et al., 2023; Singer et al., 2023).

## Conclusions

The SARS-CoV-2 pandemic made it clear that traditional event-based surveillance systems have critical deficiencies for providing prompt and valid information about the local epidemiological situation. Wastewater surveillance may provide health agencies with another early detection and effective surveillance tool, unaffected by several of the deficiencies of traditional surveillance data. As of March 2024, more than 1300 locations in the US and over 72 countries globally conducted wastewater surveillance (CDC, 2024; University of California Merced, 2024). When implemented effectively, these can data provide a comprehensive picture of SARS-CoV-2 transmission, capturing asymptomatic and non-tested infections. Our study demonstrates that analyzing wastewater metrics across multiple jurisdictions can establish the relationship between wastewater and potential cases, and how these differ across locations and over time. However, the missing data in wastewater and uncertainty in case data require future efforts to make the relationship between them more established. Efforts to collect wastewater data in a more standardized manner should be enhanced further to fully realize their potential.

## Data Availability

All data used in this study that are available online are described in the manuscript. Some data, however, cannot be shared as the authors do not have permission to do so.

## Acknowledgement

This project is supported by cooperative agreement NU38OT000297 from the Centers for Disease Control and Prevention (CDC) and the Council of State and Territorial Epidemiologists (CSTE), and SHEPheRD contract 200-2016-91779 from the CDC. This work does not necessarily represent the views of the CDC or CSTE.

