## Supplementary Material for "Statistical Relationship Between Wastewater Data and Case Notifications for COVID-19 Surveillance in the United States, 2020-2023: A Bayesian Hierarchical Model"

Appendix

### 1. Prior distribution

Prior distribution on intercepts and slopes for concentration. We use the following priors summarized in Table S-1

*Table S-1: List of prior distributions*

| Model Parameter | Distribution |
|---|---|
| Intercept term that includes a random walk effect. ($\beta_{00}$) | Normal (0, 10) |
| Coefficient of the random effect term for the intercept of the equation ($\beta_{0\_re}$) | Normal(0, 1) |
| Dispersion parameter of the random effect term for the intercept of the equation ($\sigma_{\beta 00}$ ) | Cauchy(0, 5) |
| A fixed effect of wastewater concentration ($\beta_{10}$) | Normal(0, 10) |
| Random effect of wastewater concentration ($\beta_{1\_re}$) | Normal(0, 1); |
| Dispersion parameter of a fixed effect of wastewater concentration ($\sigma_{\beta 1}$ ) | Cauchy(0, 5) |
| White noise (or random fluctuation) term for the random walk model ($\beta_{00\_fd}$) | Normal(0, sg_fd) |
| Dispersion parameter for the random walk's first differences ($\sigma_{fd}$) | Cauchy(0, 5) |
| Dispersion parameter for a negative binomial distribution ($\frac{1}{\sqrt{\varphi}}$) | Normal (0, 1) |

### 2. Figures

#### 2.1. Observed and predicted cases for all counties analyzed.

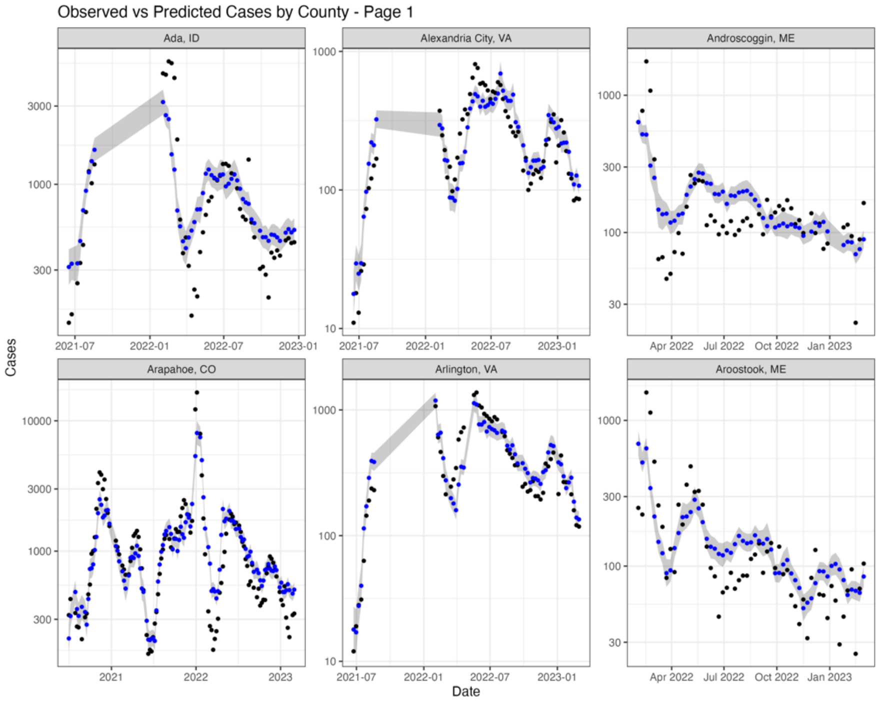

Observed vs Predicted Cases by County - Page 2

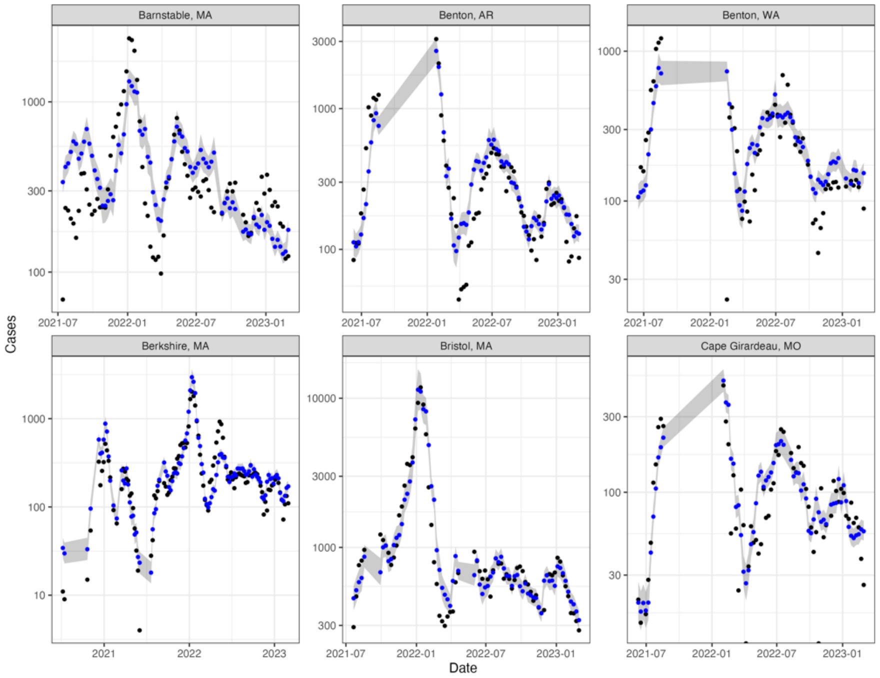


Observed vs Predicted Cases by County - Page 3

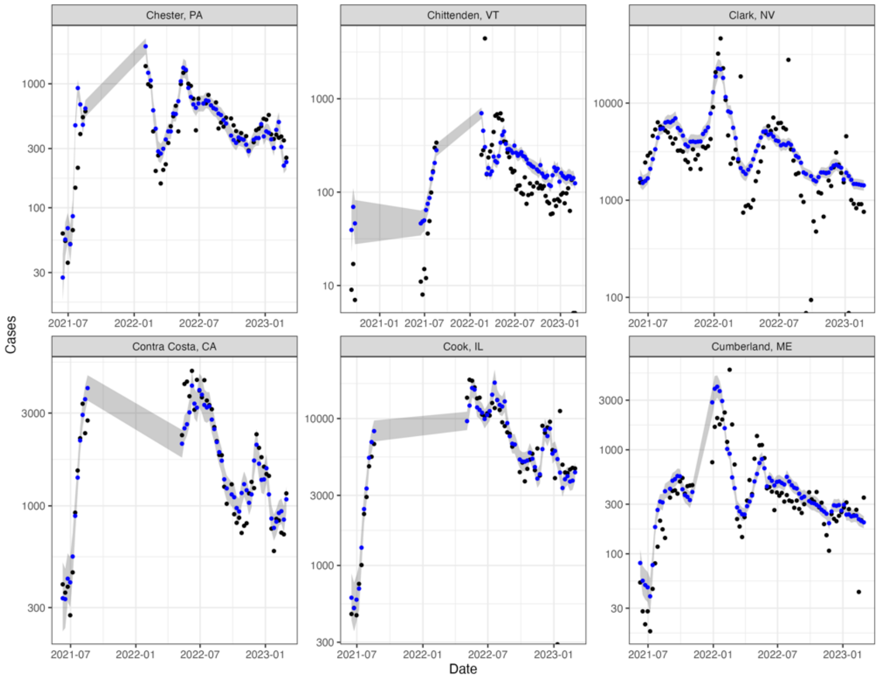

Observed vs Predicted Cases by County - Page 4

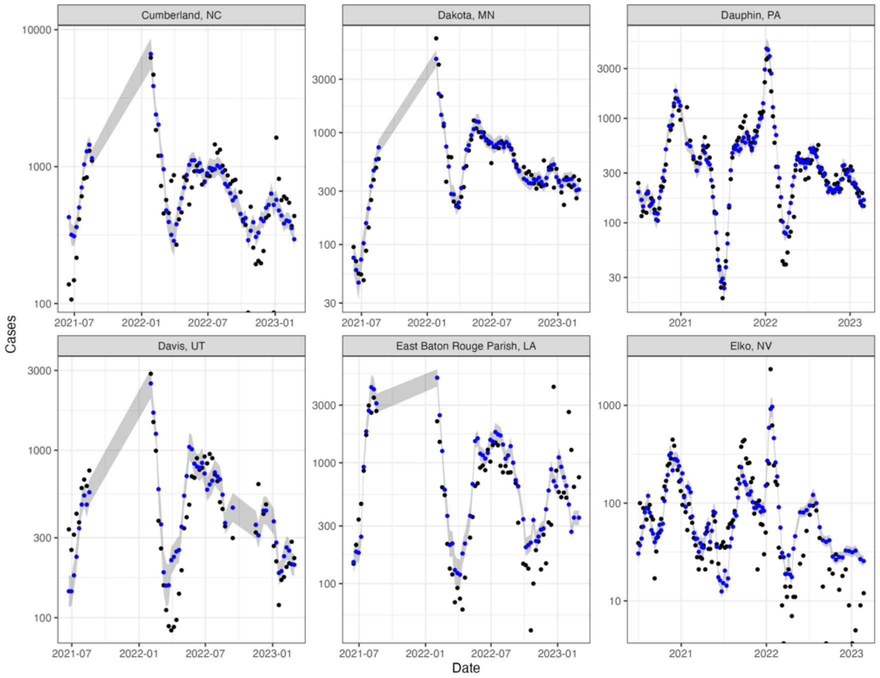


Observed vs Predicted Cases by County - Page 5

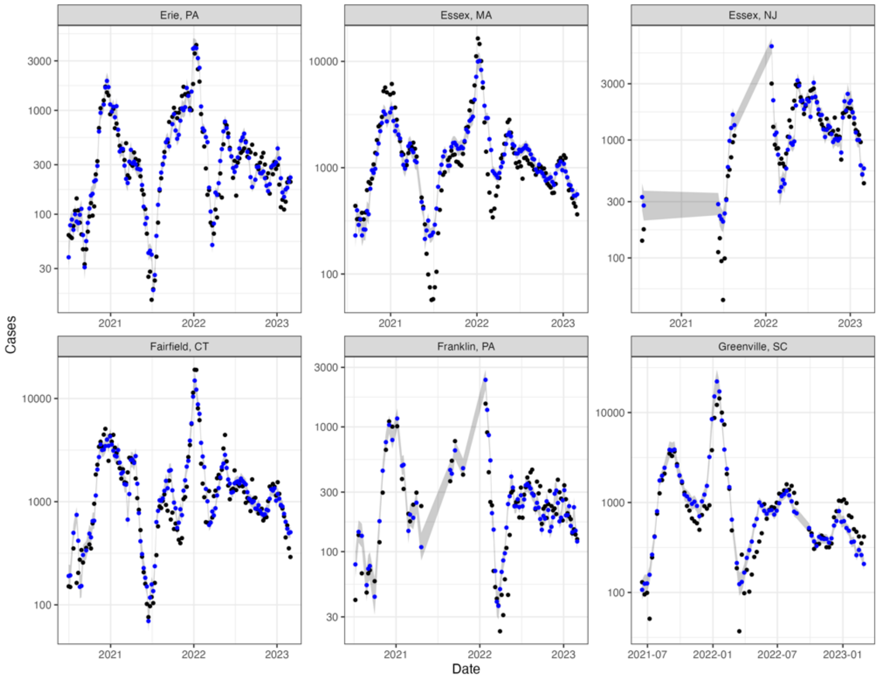

Observed vs Predicted Cases by County - Page 6

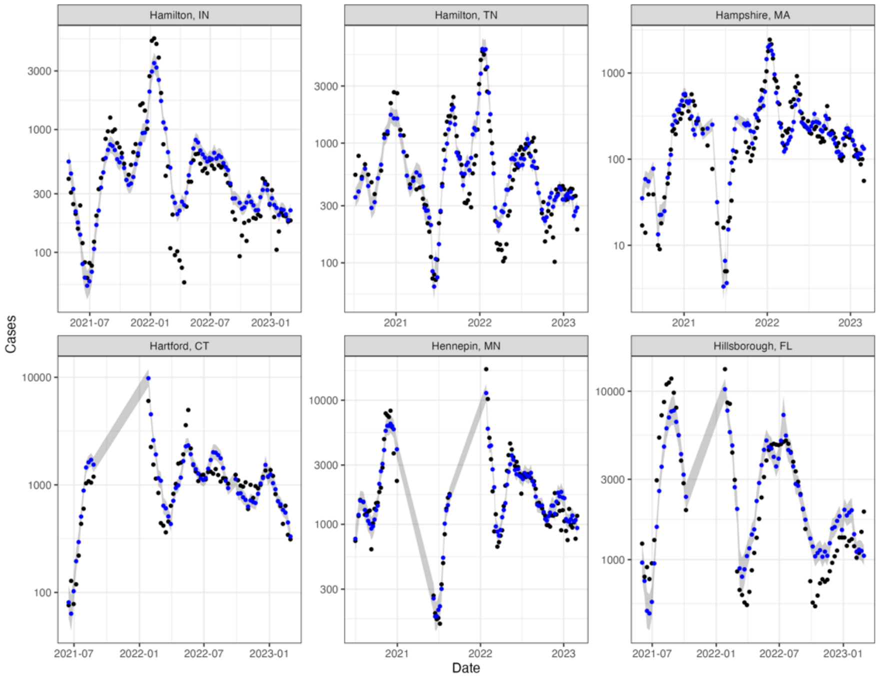


Observed vs Predicted Cases by County - Page 7

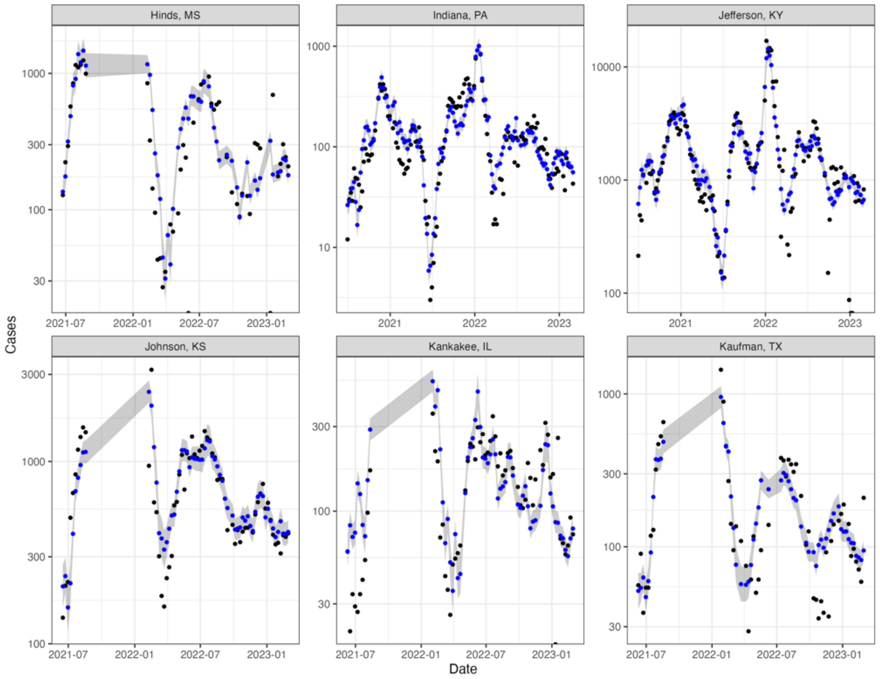

Observed vs Predicted Cases by County - Page 8

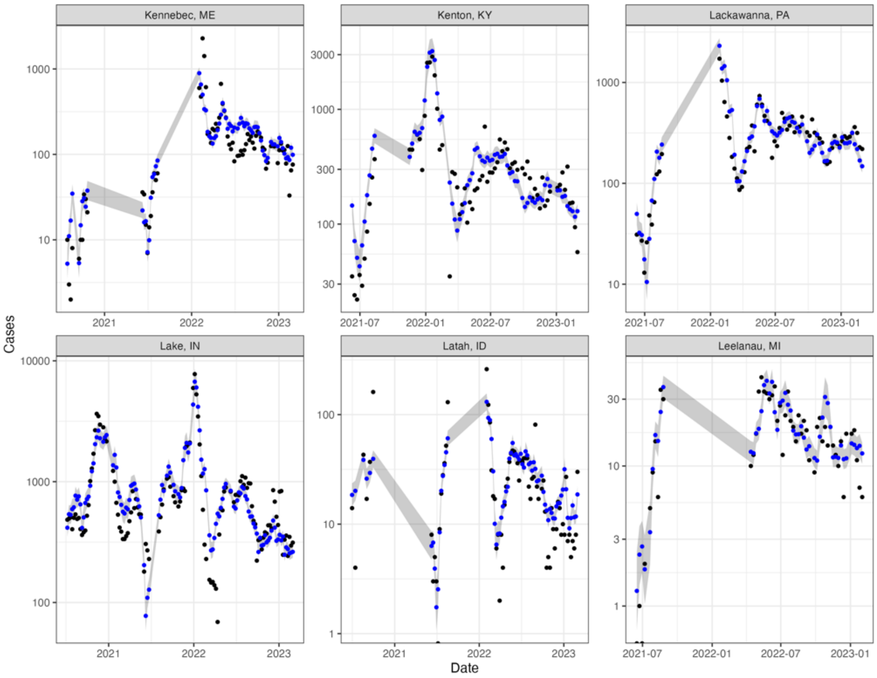


Observed vs Predicted Cases by County - Page 9

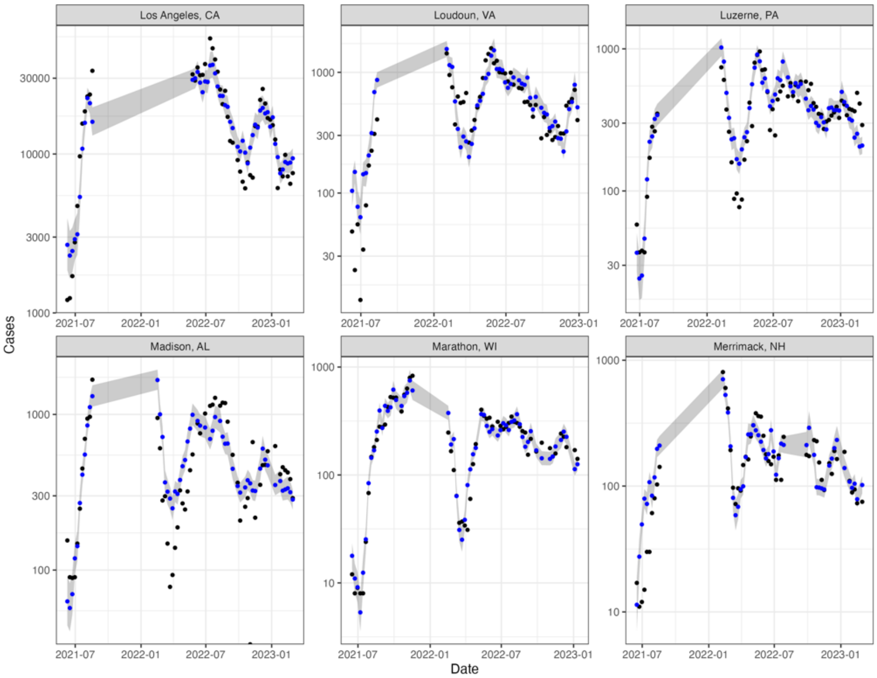

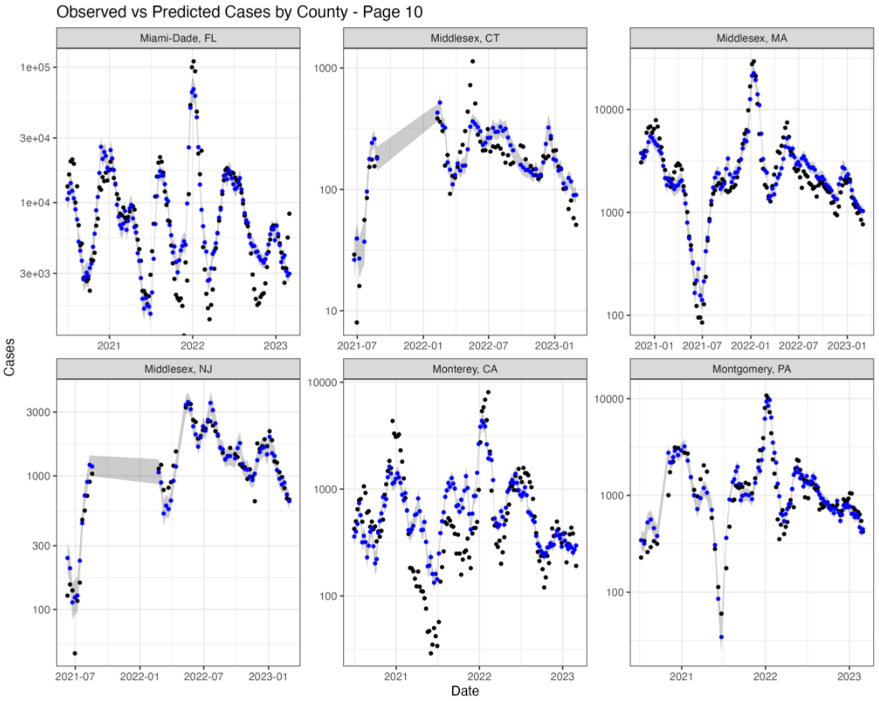
Observed vs Predicted Cases by County - Page 10
Miami-Dade, FL
Middlesex, CT
Middlesex, MA
Middlesex, NJ
Monterey, CA
Montgomery, PA
Cases
Date

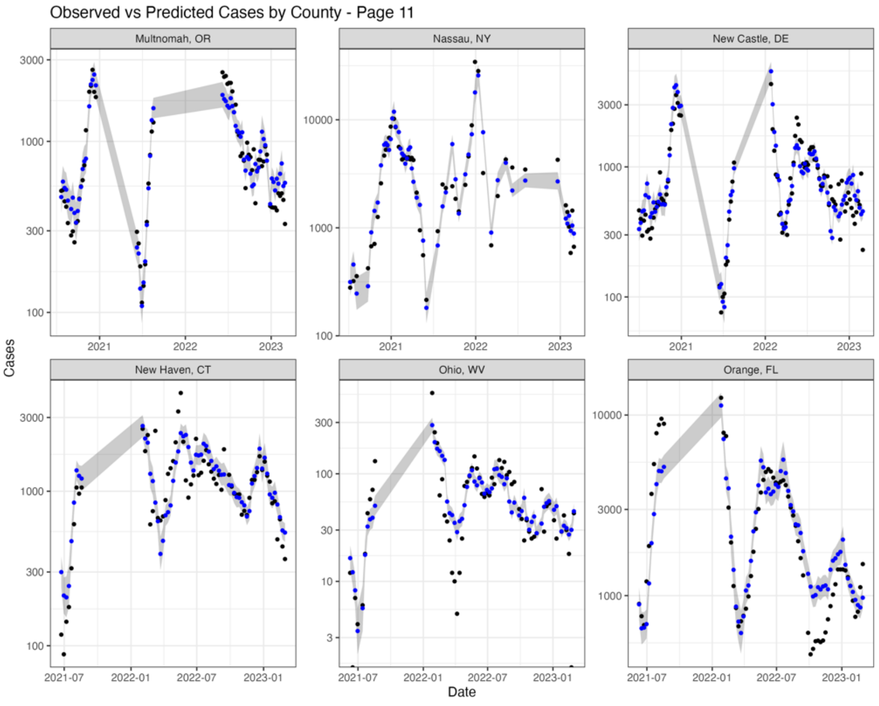
Observed vs Predicted Cases by County - Page 11
Multnomah, OR
Nassau, NY
New Castle, DE
New Haven, CT
Ohio, WV
Orange, FL
Cases
Date

#### Observed vs Predicted Cases by County - Page 12

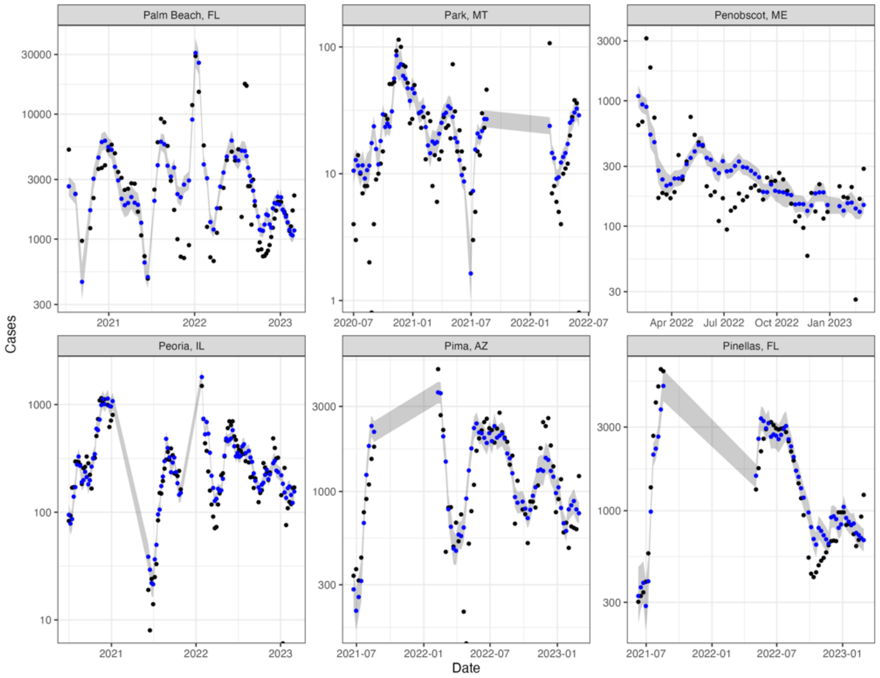


#### Observed vs Predicted Cases by County - Page 13

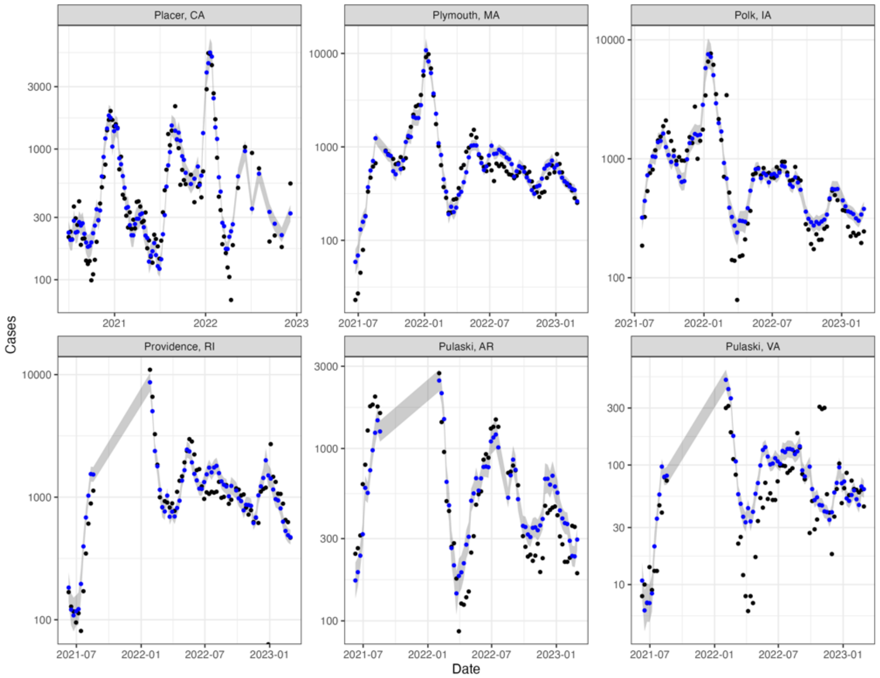

Observed vs Predicted Cases by County - Page 14

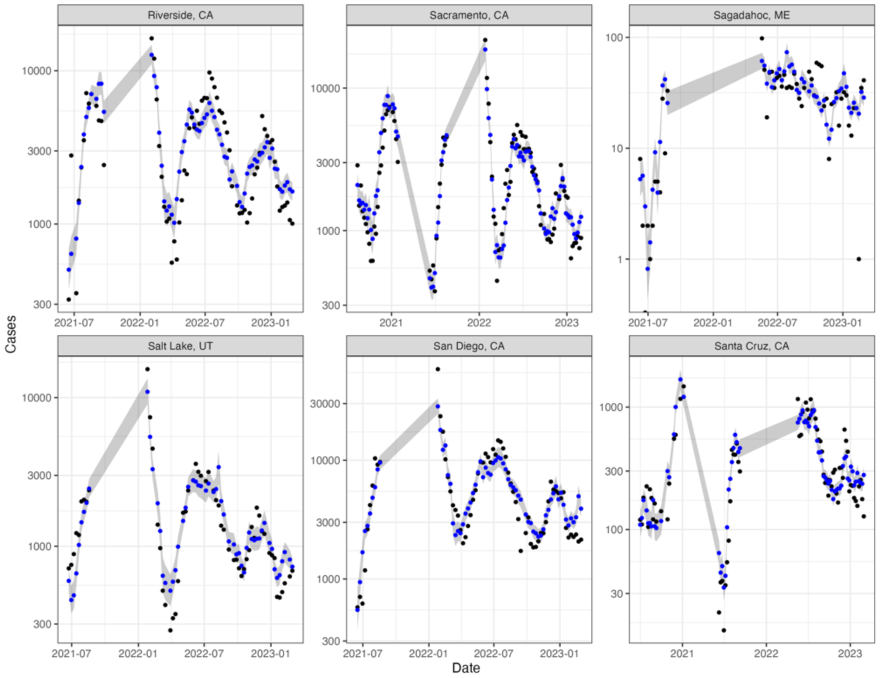


Observed vs Predicted Cases by County - Page 15

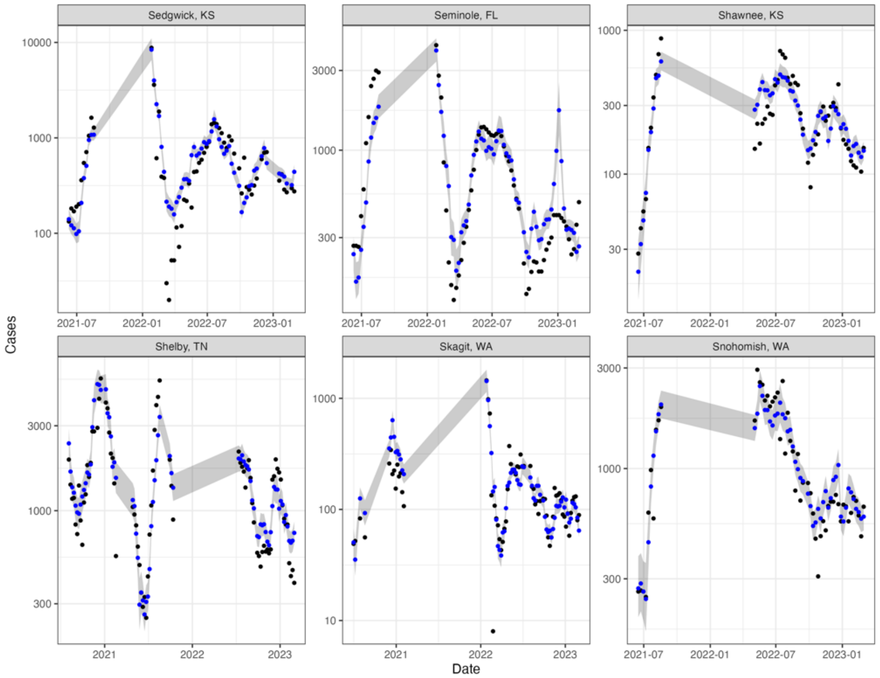

Observed vs Predicted Cases by County - Page 16

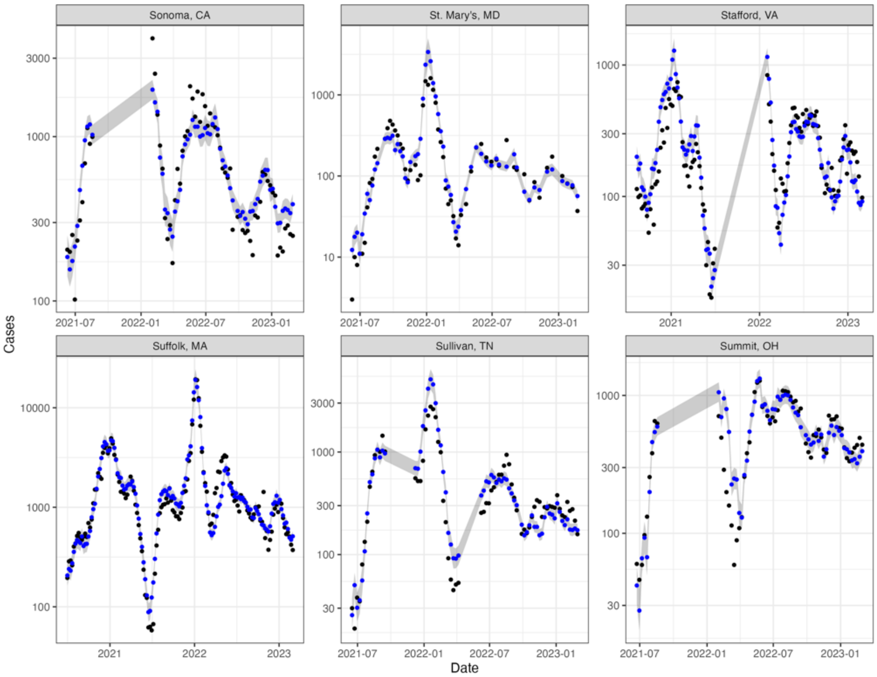


Observed vs Predicted Cases by County - Page 17

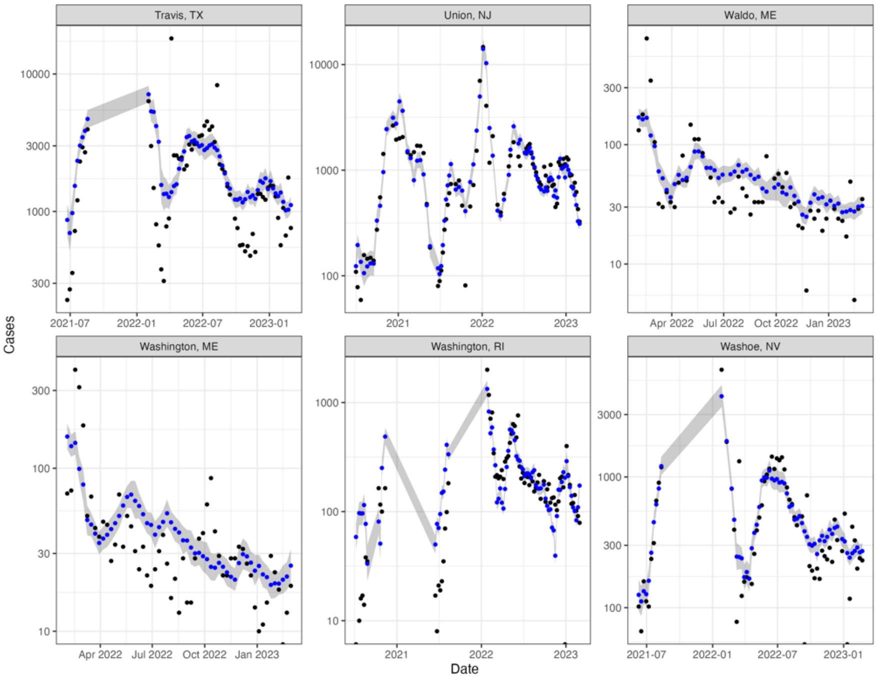

Observed vs Predicted Cases by County - Page 18

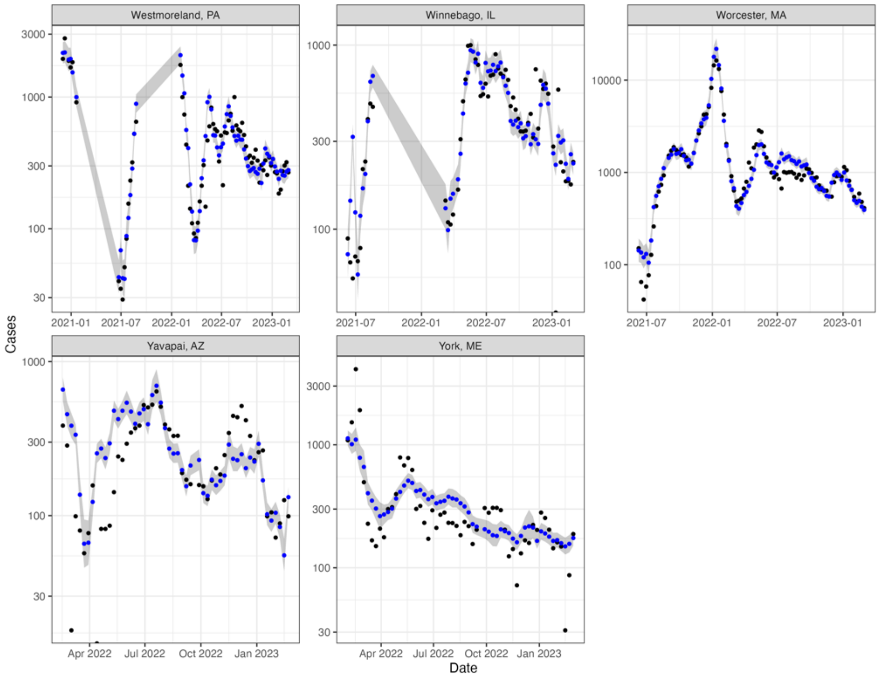
